# Understanding evolution of COVID-19 driven mortality rate

**DOI:** 10.1101/2022.01.16.22269210

**Authors:** Ishika Bhaumik, Suman Sinha-Ray, Anshul Chaudhary, Abhishek Srivastava, Prashant Kodgire

**Author notes:** Corresponding Author: Suman Sinha-Ray, University Space Research Association, NASA Glenn Research Center, Cleveland, OH, USA; /. Abhishek Srivastava, Department of Computer Science & Engineering, IIT Indore, MP, India.

## Abstract

**Objective:** COVID-19 has resulted in the death of almost 4 million people till date^1^. However, the mortality rate across countries seems to be vastly different irrespective of their respective socio-economic backgrounds. It is well known now that COVID-19 is an acute inflammatory infectious disease that gets complicated by type-I interferon response^2,3^. However, the precise reason for variations in COVID-19 related mortality rates is unknown. A detailed understanding behind the evolution of mortality rate around the globe is needed.

**Methods:** In this article, we show that a biological science guided machine learning-based approach can predict the evolution of mortality rates across countries. We collected the publicly available data of all the countries in the world with regard to the mortality rate and the relevant biological and socio-economical causes. The data was analyzed using a novel FFT driven machine learning algorithm.

**Results:** Our results demonstrate how COVID-19 related mortality rate is closely dependent on a multitude of socio-economic factors (population density, GDP per capita, global health index and population above 65 years of age), environmental (PM2.5 air pollution) and lifestyle *aka* food habits (meat consumption per capita, alcohol consumption per capita, dairy product consumption per capita and sugar consumption per capita). Interestingly, we found that individually these parameters show no visible trend that can be generalized with mortality.

**Conclusions:** We anticipate that our work will initiate conversations between health officials, policymakers and world leaders towards providing preventative measures against COVID-19 and future coronavirus-based diseases and endemics/ pandemics by taking a holistic view.

## INTRODUCTION

Coronaviruses are medium-sized RNA viruses with a characteristic crown-shaped structure. The genesis of the coronavirus dates back to 10,000 years ago^4^. However, the first known detection of coronavirus dates back to 1965^5^. While coronavirus is mainly present in bats and birds, it falls in the zoonotic category which comprises viruses that are transmitted from animals to humans from time to time. While COVID-19 seems to be extremely lethal, the surprising aspect of COVID-19 related deaths is the significant variation in mortality rates across the globe. As an example, at the height of the pandemic, the mortality rates in the USA, UK, South Korea, Kenya, and India in the month of May 2020 were of the order of 6.2%, 15.0%, 2.4%, 5.6% and 3.2%, respectively. On 2^nd^ January, 2021, the same figures were at 1.7%, 2.9%, 1.5%, 1.7% and 1.4%, respectively^1^. Researchers have put in substantial effort to explain this variation. In a recent article, it was demonstrated that the prevalence of autoimmune diseases can increase the risk of COVID-19 related fatality 1.21-1.31 folds^6^. Significant work has been done to understand the impact of hazardous air pollutants, namely: PM2.5, ozone, nitrogen dioxide^7-9^ on the fatality rates associated with COVID-19. Additionally, the effect of diet on fatality rates in European countries has also been studied. It is hypothesized that it stems from variations in angiotensin-converting enzyme activity^10^. All these endeavours clearly indicate that multiple factors affect the mortality rate associated with COVID-19. What is missing, however, is a study of the overall interdependence of various factors that influence the mortality rates associated with COVID-19. Furthermore, most studies are limited to a region and do not provide a complete picture.

In Figure 1, the mortality rate of countries across 5 different continents are elucidated, where Figure 1 a-e elucidates the mortality rate in a few countries in Africa (Nigeria, South Africa, Morocco and Kenya), Asia (India, South Korea, Japan and Singapore), Europe (UK, Italy, Germany and France), South America (Brazil, Chile, Peru and Argentina) and North America (USA, Canada and Mexico) respectively between March 15, 2020 – March 15, 2021^1^. Mortality rate (called as Case Fatality Rate in Ref. 1) is defined as the following-

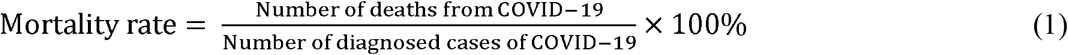

**Fig. 1.**
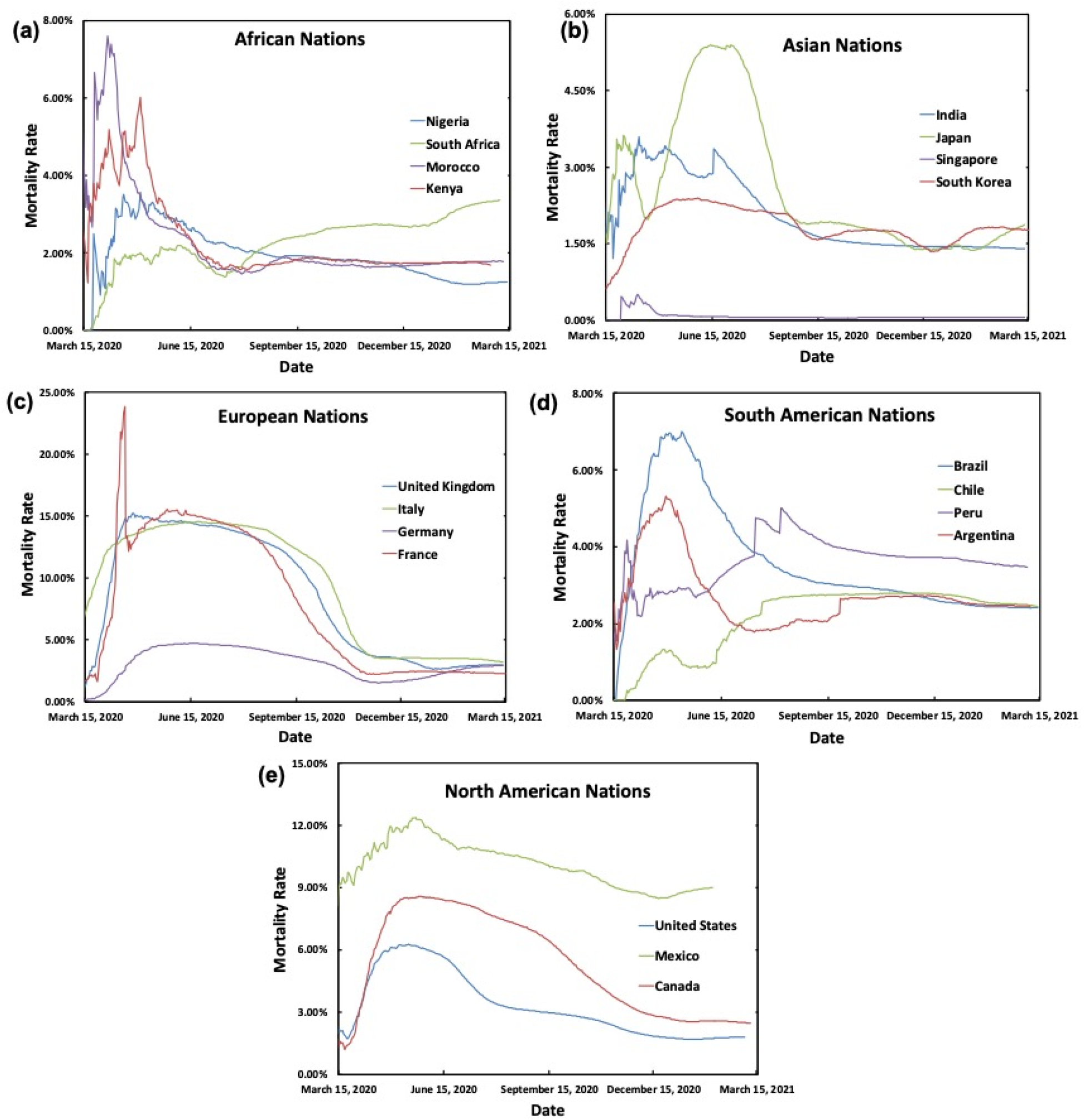
The variation of mortality rate across nations in various continents over 1 year period (March 15, 2020-March 15, 2021) are shown – (a) African (Nigeria, South Africa, Morocco and Kenya), (b) Asian (India, Japan, South Korea and Singapore), (c) European (UK, Italy, Germany and France), (d) South American (Brazil, Chile, Peru and Argentina) and (e) North American (USA, Canada and Mexico) nations.

While such definition has its inherent challenges^1^, we believe that this is the best way to normalize the data and compare between countries of different sizes and populations. Figure 1 brings in two interesting facts. First, as can be seen, almost all the countries follow a very similar trend in the evolution of mortality rates across time. While mortality rates during the initial days were the highest, they subsequently went down and plateaued with a significant exception in South Africa, which perhaps can be attributed to the surge of the Beta (501Y.V2) variant^11^. This simple graph clearly shows that during the early days of the pandemic, as there was a significant lack of knowledge base on the coronavirus, the mortality rate jumped significantly. As the knowledge grew, best practices (e.g.-hygiene and sanitation, vaccinations, travel restrictions) were practiced throughout the globe and the mortality rate dropped. However, this also resulted in countrywide lockdowns resulting in economic hardships across the globe. This also hints that in future for any other novel coronavirus related pandemic, the mortality rate can be similarly higher. Secondly, it can also be seen that the mortality rates across different countries are largely different. The most interesting aspect is that the mortality rates in several rich nations are higher than their poorer counterparts. All these beg the question as to the reason behind the driver of the mortality rate. It can be seen that owing to the variation of exposure rate, the COVID-19 related mortality rate peaked at different times. Additionally, it can also be seen that for some countries, the mortality rate started dropping in early 2021. However, that drop was not uniform across the world. This can be attributed to the uneven distribution of vaccines across the globe. To avoid these variations and spikes, our analysis was limited to the period between 15^th^ May 2020 and 15^th^ January 2021.

## MATERIALS AND METHODS

### Determination of the Influencing Factors

As COVID-19 is an inflammatory disease^2,3^, our primary hypothesis at the beginning of this work was that any factor that creates an inflammatory response in the human body will be accentuated by COVID-19 resulting in an increased mortality rate. The variation of the impact of COVID-19 on the mortality rate can be ascribed to long term influencers that create heightened inflammation in the body. In addition to that, recently there has been some interesting work that clearly show that COVID-19 is severely correlated to gut microbiome^12^. While choosing the factors, our hypothesis also evolved around the fact that if the factors also negatively impact the gut microbiome, it will cause severity of complications by COVID-19 and thus increase the mortality rate. It also needs to be mentioned that our approach was to utilize the minimum number of influencing factors to correlate with the mortality rate.

One of the most established factors in COVID-19 related deaths is the ageing population^13^. While that is a significant factor, it is worth noting that as of 2020 while France has 20.75% of the population with an age of 65+, Japan has 28.4% of their population with an age of 65+^1^. The mortality rate plot in Figures 1b and 1c clearly shows that France had a significantly higher mortality rate than Japan. PM 2.5 also has a significant impact on the mortality rate associated with COVID-19^7-9^. Such a dependence is possible because as it was found that PM 2.5 can be associated with enhanced inflammatory markers resulting in diabetes, obesity, hypertension prevalence in patients^14-17^. Another popular aspect to the discrepancy in mortality rate was ascribed to access to health care, sanitation, hygiene, higher population density and GDP. Better hygiene and safe sanitation practices in high GDP countries are also associated with a higher prevalence of autoimmune disorders, diabetes *aka* higher inflammatory response^18^.

Additionally, high GDP results in a sedentary lifestyle causing a multitude of health issues. One of the most controversial yet significant reasons behind enhanced inflammatory response in the human population are dietary habits^19,20^. These studies characterized the role of food with saturated fat and milk with disorders like Multiple Sclerosis (MS) and diabetes. Interestingly, this study reported that whole food, plant-based or high fiber diets help in controlling diabetes-related complications and consumption of red meat developed evidence of systemic inflammation and thereby contributed to a higher incidence of type 2 diabetes and atherosclerosis. Similarly, heavy alcohol consumption demonstrated higher C-reactive protein (CRP) concentrations that is an important systemic marker of inflammation^21^. With regard to the correlation between the gut microbiome and the above-mentioned factors, it was found that the factors not only negatively impact inflammation, they also negatively impact the gut microbiome ^22-26^. Based on all these, we hypothesized that long term exposure of the following - socio-economic factors (population density, GDP per capita, global health index and population above 65 years of age), environmental (PM2.5 air pollution) and lifestyle *aka* food habits (meat consumption per capita, alcohol consumption per capita, dairy product consumption per capita and sugar consumption per capita), drives the mortality rate.

### Overview of the Scientific Model

A schematic explaining the overview of the scientific model is shown in Figure 2. The overall evolution of mortality rate (Figure 1) for all the countries was made to undergo an FFT (Fast Fourier Transformation) yielding the Fourier coefficients (a_n_’ s and b_n_’ s). Machine learning-based models were trained to correlate the Fourier coefficients with the above-mentioned influencing factors. From the training data set, Fourier coefficients of the validation dataset were predicted, and these underwent an IFFT (Inverse Fast Fourier Transformation) to predict the mortality rate. The mortality rates so predicted were compared with the actual mortality rates and the efficacy of the approach was assessed. To the best of our knowledge, this process of solving an epidemiological model is novel. This idea of solving complex evolution in the Fourier space is more common in understanding turbulence^27^ and the frequency of oscillation of a heating wire in boiling heat transfer^28^. This method is one of the novel aspects of our present work and is described in detail in the following subsections.

**Figure 2.**
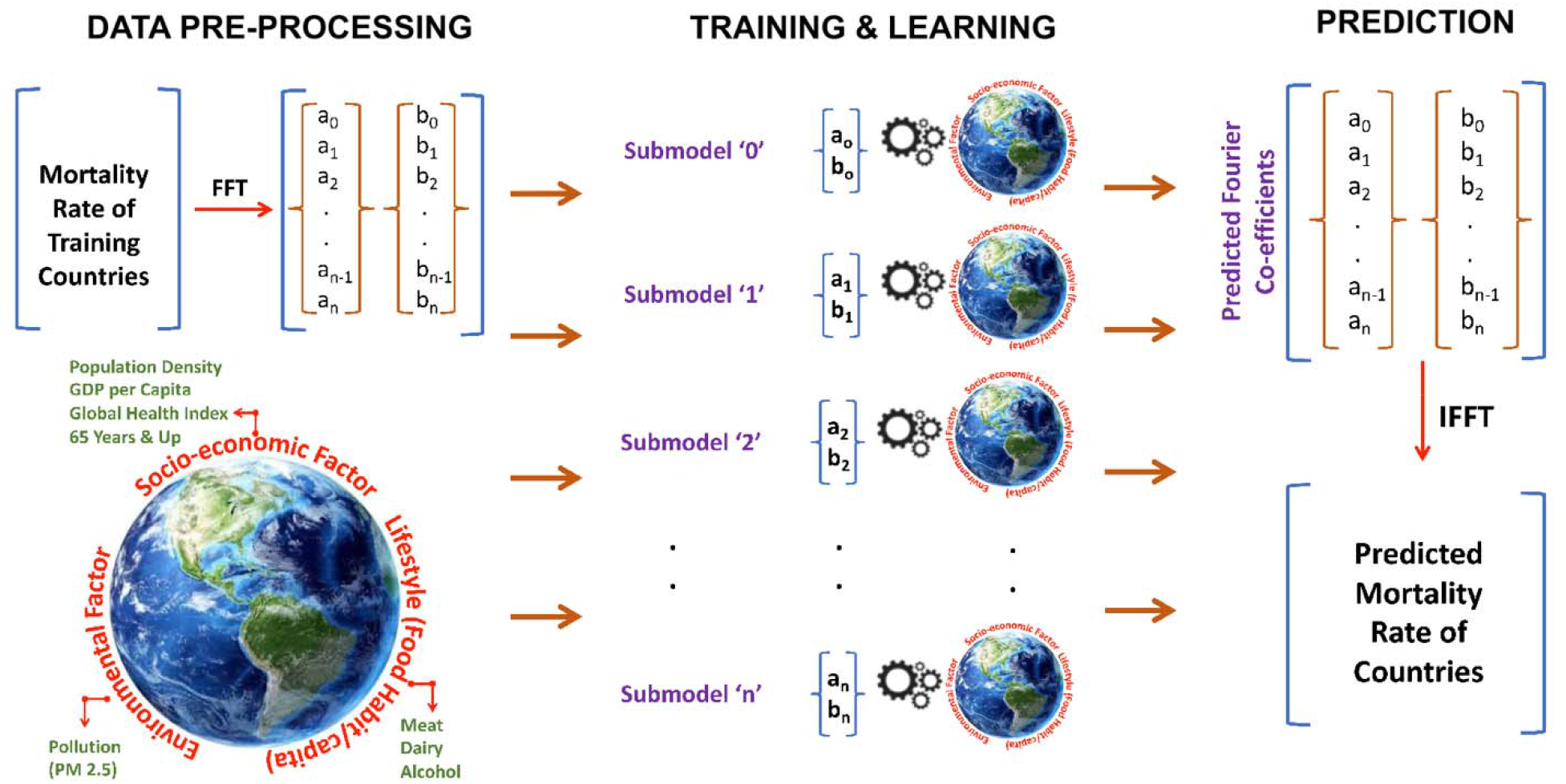
An overview of the scientific model is shown. The model has 3 distinct parts-data pre-processing, training & learning and prediction.

### Data Collection

The influencing factors were obtained from publicly available data from reliable sources^1^. While these data provide an overall view of a country, it lacks the granular details. This lack of granularity may cause some variation in the accuracy of prediction. However, our overarching hypothesis was that such general data will still be able to provide the predictive capability. The data for the number of COVID-19 cases, number of deaths for the aforementioned period is collected for almost all countries in the world. In addition to this, data corresponding to the parameters mentioned is collected for all countries for the last ten years. Algorithm 1 summarises our data collection efforts. The number of covid cases per million for each country and the number of deaths due to covid per million for each country is calculated defined as Algorithm 1 is shown in Figure 3a.

**Figure 3.**
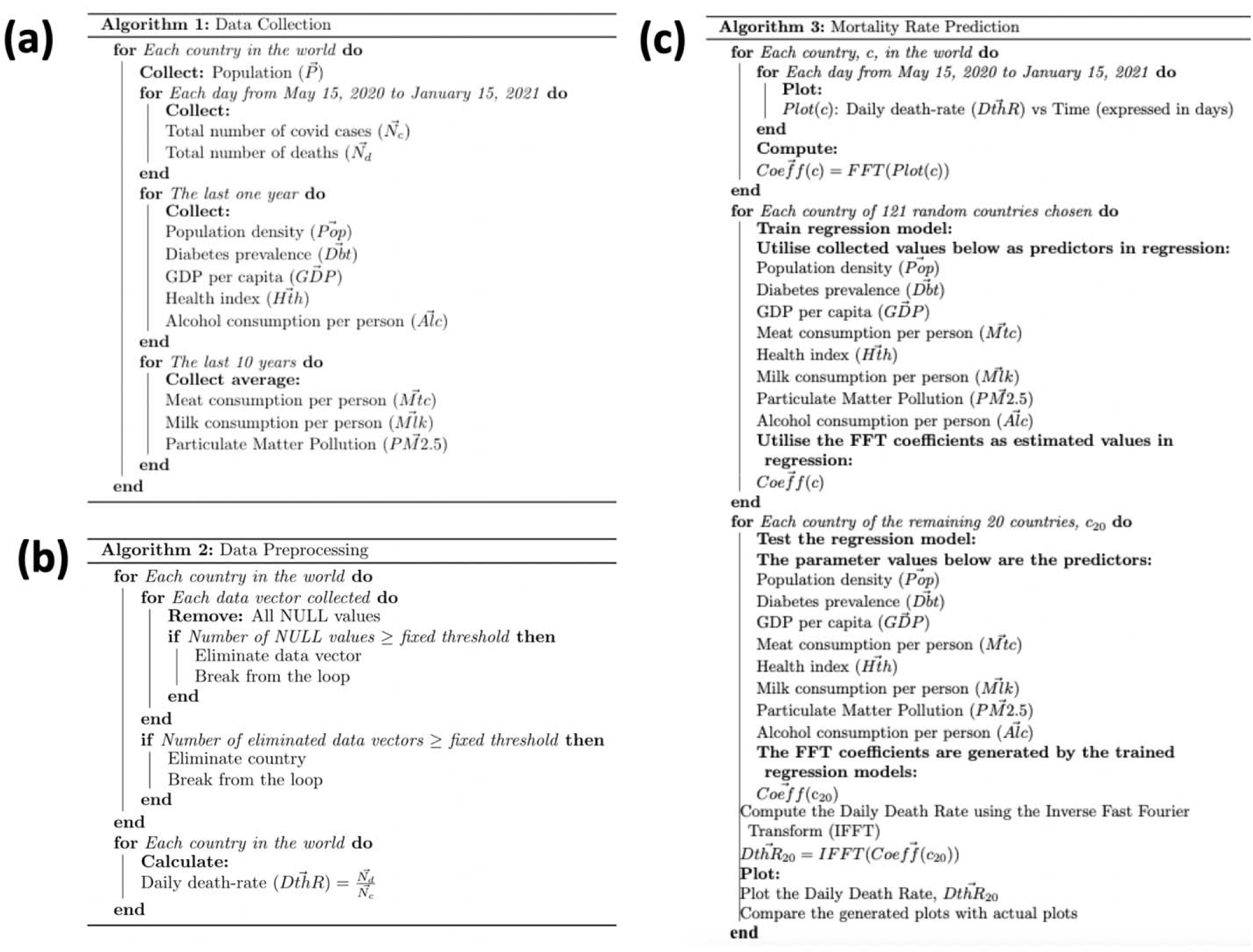
(a) Data collection methodology is shown (Algorithm 1). (b) Data pre-processing methodology is shown (Algorithm 2). (c) The machine learning model is shown (Algorithm 3).

### Data Pre-processing

While the publicly available data is comprehensive, in some cases it lacked the details. In order to utilize the ‘dirty’ data, it need to be ‘cleaned up’ first to be effective in providing useful information. We utilised the Pandas software library for proper consolidation of data. The data is mostly available in a scattered manner in a CSV format. We consolidated data for each of the parameters in such a way that all data for a specific parameter for different countries is gathered together. In case the data corresponding to a certain country is missing or is NULL, the parameter value is eliminated. In case the number of NULL values corresponding to a country exceeds a certain threshold, the country is eliminated and is not considered for the analysis. The data for the parameters in a country-wise manner is put together as a data frame. Subsequent to the elimination of the NULL values and/or countries with a large number of NULL values for their respective parameters, the death rate for each country is calculated as the ratio of the number of deaths per million to the number of COVID-19 cases per million of the respective country. Algorithm 2 summarizing the data pre-processing is shown in Figure 3b.

### Development of the Machine Learning Model

In this work, we focused on predicting the mortality rate trend between 15^th^ May 2020-14^th^ February, 2021. Instead of looking at daily fluctuation, which can stem from a multitude of nation-specific causes, the overall long-term trend provides a holistic view. The above-mentioned pre-processing step resulted in 141 countries. Of these, the machine learning model was trained on 121 and rest 20 were chosen as the validation set. From Fig. 1 of the main text, it can be seen that the evolution of the mortality rate is random. In order to explain the evolution in a mathematically closed-form solution, FFT (Fast Fourier Transformation) was used. In FFT, the functional representation of the evolution can be explained as

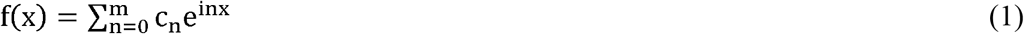

, where c_n_ = a_n_ + ib_n_, where a_n_ and b_n_ are Fourier coefficients and m is the number of days. Then the influencing factor(s) of the training dataset were trained against the Fourier Coefficients of the corresponding training data using gradient boosting regressor with a learning rate of 0.001 and a value of n_estimators (number of regression trees) as 100. For ‘m’ Fourier coefficients, ‘m’ submodels were employed to train the algorithm. Subsequent to training the regressor with data for 121 countries, the same is utilised to predict the FFT coefficients for the 20 countries in the test set using the respective parameter values. The Fourier coefficients so generated are subsequently made to go through an Inverse Fast Fourier Transformation (IFFT) yielding the mortality rate of the countries in the training dataset, which was then compared with the actual mortality rate data. Algorithm 3 summarizing our approach for the prediction of the mortality rates is shown in Figure 3b.

## RESULTS AND DISCUSSION

### Prediction of Mortality Rate Using Single Parameter

Using the above-mentioned protocol, as shown in Figures 2 and 3, investigations were first conducted to predict/ explain the trend in COVID-19 related mortality rates incorporating just one of the influencing factors. A comparison between the USA and Slovenia is shown in Figure 4. Some of the other countries’ data is shown in Figure S1-S10. It is clear that a single influencing factor cannot explain the mortality rates across countries. Figure 4 shows that for Slovenia, alcohol consumption and global health index seem to predict the mortality rates close to the actual data (Figure 4a and 4d), for the USA there seems to be no trend for these two factors. For the USA it seems that GDP per capita and PM 2.5 (Figure 4c and 4g) closely predict the mortality rate. This is one of the challenges we found with the results in existing literature, where correlations were drawn using a small subset of data. In order to alleviate this, the protocol shown in Figures 2 and 3 were followed and a comprehensive model utilizing all the factors together was developed.

**Figure 4.**
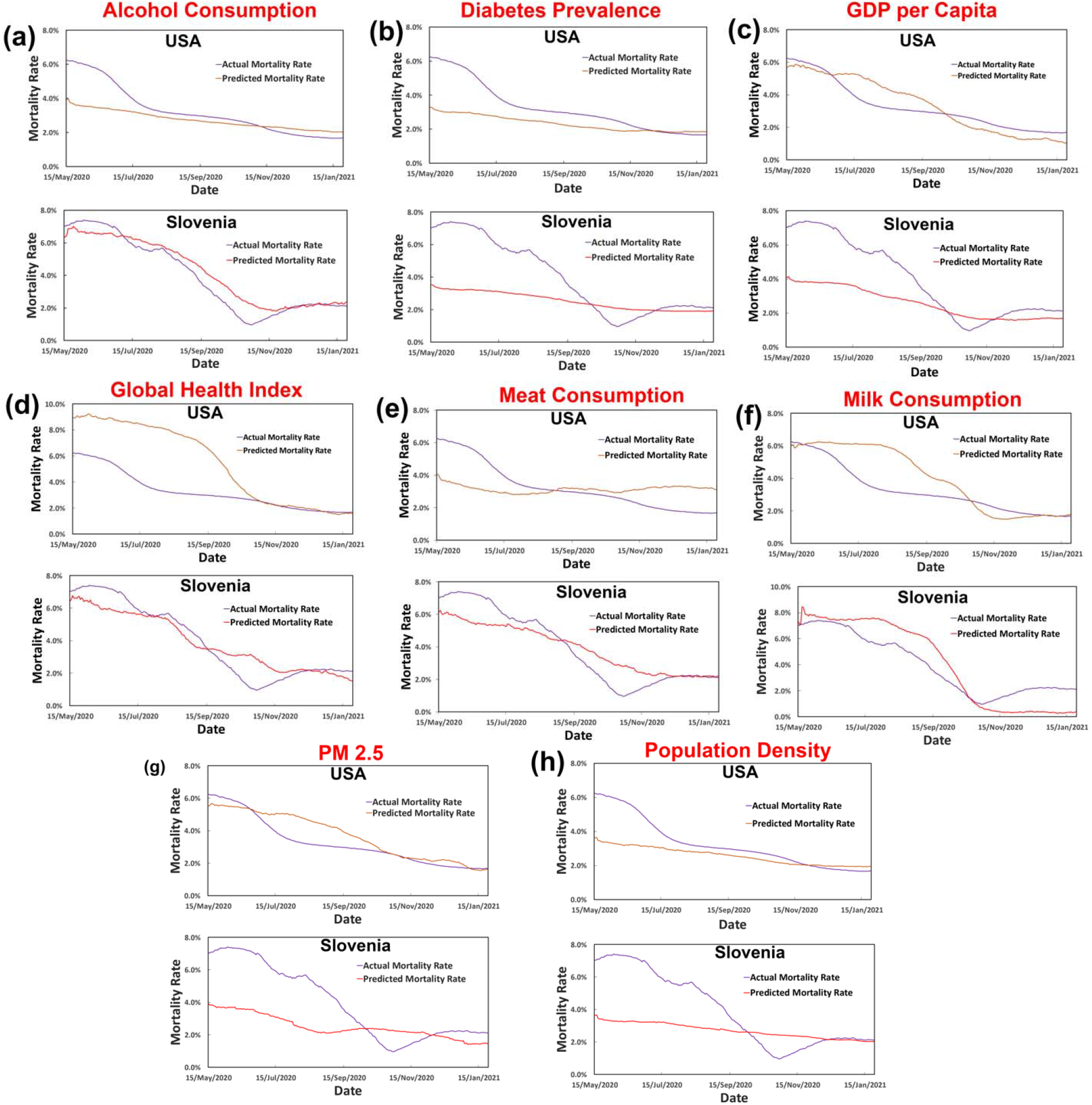
Prediction of COVID-19 related mortality rate using a single factor for both USA and Slovenia are shown, where the factors are – (a) alcohol consumption, (b) diabetes prevalence, (c) GDP per capita, (d) global health index, (e) meat consumption, (f) milk consumption, (g) PM 2.5 and (h) population density. None of the factors individually can describe the trend comprehensively.

### Prediction of Mortality Rate Using 8 Crucial Parameters

Using the proposed protocol (Figures 2 and 3), the mortality rates of 20 randomly selected countries were predicted and compared with the actual mortality rates. Results of 6 of the countries are shown in Figure 5 of the main text and the results of some of the remaining countries are shown in Figure S11 in the supplementary information. Figure 5a-f corresponds to the USA, Germany, Honduras, Slovenia, Togo, and Brazil respectively. It is clear that for all the countries, the predicted mortality rates are very close to the actual mortality rates. The variations that do exist stem from multiple factors-(a) the quality of COVID data has challenges; (b) the publicly available data used in the prediction lacks granularity and specificity; and (c) the physiological difference (e.g.-blood type) may also potentially result in significant variations in mortality rate^29,30^. Such granular information is practically impossible to obtain for the world population. As a result of which the prediction has an ingrained challenge in accurately predicting the mortality rates. However, the interesting and heartening fact is that irrespective of the paucity of granular data, our predictions not only show the impact of the 8 influencing factors on mortality rates but are also able to precisely trace the evolution and trend of the mortality rates across the world population. This clearly holds potential to positively impact the preparations for future coronavirus related outbreaks. The large message is that policymakers, government officials and scientists need to take a holistic approach towards the healthcare response.

**Figure 5.**
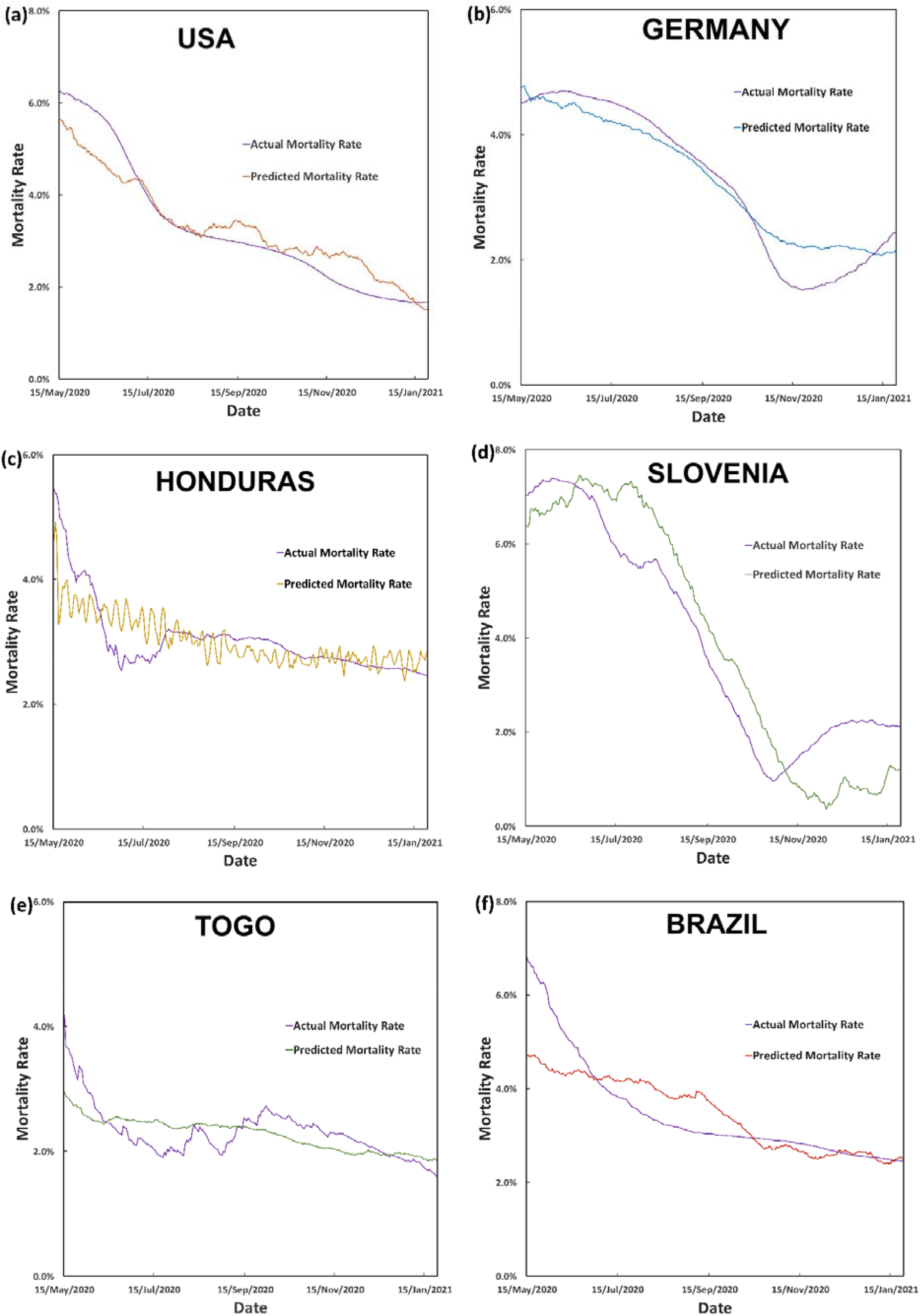
Prediction of COVID-19 related mortality rate using 8 factors and their comparison with actual mortality rates are shown for –(a) USA, (b) Germany, (c) Honduras, (d) Slovenia, (e) Togo and (f) Brazil are shown. It can be seen that the model is capable of predicting the evolution of mortality rate quite well.

## CONCLUSIONS

This work utilizes a novel FFT guided machine learning methodology to demonstrate that COVID-19 related mortality can be broadly related to multitude of socio-economic factors (population density, GDP per capita, global health index and population above 65 years of age), environmental (PM2.5 air pollution) and lifestyle *aka* food habits (meat consumption per capita, alcohol consumption per capita, dairy product consumption per capita and sugar consumption per capita). Owing to the lack of granularity of individual level data and different country level introduction of security protocol during pandemic (e.g.-imposition of lockdown etc.), the prediction may not exactly fit to the actual mortality rate data but it closely elucidates the trend. This clearly shows that the policymakers, health professionals and government need to take a long-term view to protect against any future coronavirus-mediated pandemic. Also, we believe that further study building onto this one will also allow the policymakers to take actions more proactively in future.

## Data Availability

All data produced in the present study are available upon reasonable request to the authors

## AUTHOR CONTRIBUTIONS

I.B. carried out the model development. S.S.R. conceptualized the idea and directed the research. A.C. collected and prepared the data for analysis. A.S. directed the research. P.K. contributed to identifying the biological significance of the factors. All authors contributed to the analysis of the results and the writing of the manuscript.

## SUPPLEMENTARY MATERIAL

Supplementary material is available at Journal of the American Medical Informatics Association online.

